# Overweight followed by obesity have the lowest mortality among patients with systolic or diastolic heart failure independent of comorbidities whereas cachexia has the highest mortality

**DOI:** 10.1101/2024.01.17.24300813

**Authors:** Mohammad Reza Movahed, Austin Mineer, Mehrtash Hashemzadeh

## Abstract

**Introduction:** A phenomenon known as the obesity paradox has been reported in patients with heart failure (HF) suggesting lower mortality with increasing weight. The goal of this study is to characterize this observation in HF using the largest available inpatient database of adult patients.

**Method:** We searched the National Inpatient Sample (NIS) database for patients for the years 2016-2020 with a diagnosis of systolic (SHF) or diastolic heart failure (DHF) using International Classification of Diseases, Tenth Revision (ICD-10) diagnosis codes. We evaluated mortality association based on body weight. Multivariate statistical analysis was performed to adjust all-cause inpatient mortality for comorbidities.

**Results:** There was a total of 7,364,023 patients with a diagnosis of SHF and 10,064,223 patients with a diagnosis of DHF. All-cause inpatient mortality was lowest in overweight patients followed by those with obesity and morbid obesity, whereas mortality was highest in cachexia compared to normal weight for SHF and DHF patients (mortality: overweight 2.56%, obese 3.12%, morbidly obese 3.70%, normal weight 5.60%, and cachexia 15.22%; p<0.001) and DHF patients (mortality: overweight 2.08%, obese 2.43%, morbidly obese 2.93%, normal weight 4.58%, and cachexia 14.25%; p<0.001). This relationship remains similar after multivariate analysis. (SHF patients: overweight OR: 0.49 (0.41-0.58), obesity OR: 0.64 (0.62-0.66), morbid obesity OR: 0.85 (0.83-0.88), and cachexia OR: 2.78 (2.67-2.90); p<0.001; DHF patients: overweight OR: 0.47 (0.40-0.56), obesity OR: 0.61 (0.59-0.63), morbid obesity OR: 0.83 (0.81-0.85), and cachexia OR: 3.09 (2.96-3.23); p<0.001).

**Conclusion:** Our data observed that all-cause inpatient mortality in SHF and DHF is lowest in overweight populations followed by obese and morbidly obese populations whereas cachexia has the highest mortality. However, increasing weight above the overweight reduces the obesity paradox benefit.

## Introduction

The incidence and prevalence of obesity has risen dramatically in the United States and has reached pandemic levels worldwide.^1,2^ The disease has been extensively studied and excess visceral adiposity has been shown to impose a variety of systemic metabolic derangements, such as increasing pro-inflammatory signaling cascades, renin-angiotensin-aldosterone system activation, insulin resistance, and blood vessel sympathoactivation,^3,4^ leading to many chronic diseases. Particularly, it serves as a significant risk factor in the pathogenesis of cardiovascular disease (CVD),^3,4,5,6^ including hypertension, coronary heart disease, atrial fibrillation, and heart failure (HF).

Still, our collective understanding of weight and obesity continues to grow. Although it continues to serve as a major comorbidity in the development of CVD, studies have shown that in patients with established CVD, overweight and obese populations display a better prognosis in comparison to those with normal weight.^5,6,7,8,9^ This “obesity paradox” in CVD is especially apparent in HF, although questions regarding this phenomenon persist and warrant further examination. Some have suggested that in patients with HF, mortality decreases with increasing weight,^8,9,10,11,12^ whereas others have proposed that there is a specific point where the detrimental effects of obesity are minimized and the beneficial effects of the obesity paradox are maximized.^5,6,7,13^ Still, results are unclear, and the point of weight optimization remains uncertain. Furthermore, due to obesity’s complex interplay with various metabolic systems, comorbidities of obesity make studying its effects on HF challenging and bring uncertainty to our current understanding of the relationship. The goal of this study is to evaluate and characterize the obesity paradox in both SHF and DHF patients with the largest available inpatient database of adult patients while eliminating potential confounders by adjusting for baseline characteristics and comorbidities of patients.

## Method

### Collected Information

This study contains data from patients aged over 18 years old that were admitted to NIS hospitals from 2016 to 2020, appointed an ICD-10 diagnosis code for SHF (I50.2, I50.20, I50.21, I50.22, I50.23) or DHF (I50.3, I50.30, I50.31, I50.32, and I50.33). These patients were categorized according to ICD-10 diagnosis codes used for the following body weights: cachexia (R64), overweight (E66.3), obesity (E66.9, E66.8, E66.0), and morbid obesity (E66.01, E66.2). These ICD-10 codes correspond to different body mass indexes (BMI) and are defined as follows: <18.5 is considered underweight (cachectic), 18.5 to <25.0 is considered normal weight, 25.0 to <30.0 is considered overweight, 30.0 to <40.0 is considered obese, and >40.0 is considered morbidly obese. Comorbidity data was collected on these patients also using ICD-10 diagnosis codes (Table 1) for the following comorbidities of HF: diabetes, hypertension, chronic obstructive pulmonary disease (COPD), chronic kidney disease (CKD), ST elevation myocardial infarction (STEMI), non-ST elevation myocardial infarction (NSTEMI), and old myocardial infarction (MI). Patient demographics and details of their hospital visit were additionally collected including age, length of stay, total charges, sex, race, insurance, hospital bed size, ownership of hospital, hospital location and region, and median household income (Tables 2, 3).

**Table 1:** ICD-10 diagnosis codes used in searching the NIS database (COPD = chronic obstructive pulmonary disease, CKD = chronic kidney disease, MI = myocardial infarction, STEMI = ST elevation myocardial infarction, NSTEMI = non-ST elevation myocardial infarction).

|  |  |
| --- | --- |
| SHF | I50.2, I50.20-I50.23 |
| DHF | I50.3, I50.30-I50.33 |
| Cachexia | R64 |
| Overweight | E66.3 |
| Obesity | E66.9, E66.8, E66.0 |
| Morbid Obesity | E66.01, E66.2 |
| Smoking | F17.20, Z72.0, Z87.891 |
| Diabetes | E08-E13 |
| Hypertension | I10, I11.0, I11.9, I120, I129, I13.0, I13.10, I13.11, I13.2, I15.0, I15.1, I15.2, I15.9, I16.0, I16.1, I16.9 |
| COPD | J41.0, J41.1, J41.8, J42, J43.0, J43.1, J43.2, J43.8, J43.9, J44.0, J44.1, J44.9, J47.0, J47.1, J47.9, J684 |
| CKD | I13.11, I13.2, N289, Q613, N181, N182, N183, N1830, N1831, N1832, N184, N185, N186, N189, R880, N19 |
| STEMI | I21.01, I21.02, I21.09, I21.11, I21.19, I21.21, I21.29, I21.3, I21.9, I21.A1, I21.A9, I22.0, I22.1, I22.5, I22.9 |
| NSTEMI | I21.4, I22.2 |
| Old MI | I25.2 |

### Statistical Analysis

Patient demographic, clinical, and hospital characteristics are means with standard deviation for continuous variables and proportions, and 95% confidence intervals for categorical variables. Multivariable logistic regression was performed for variable significance in univariate analysis to adjust the odds of clinical outcomes relative to patient and hospital characteristics as well as ascertain the odds of clinical outcomes over time. All analyses are conducted following the implementation of population discharge weights. All p-values are 2-sided and p<0.05 is considered statistically significant. Data was analyzed using STATA 17 (Stata Corporation, College Station, TX).

## Results

There was a total of 7,364,023 patients with a diagnosis of SHF that were stratified by weight groups and demographics, with a total all-cause inpatient mortality rate of 5.34% (Table 2). There was a total of 10,064,223 patients with a diagnosis of DHF that were stratified by weight groups and demographics, with a total all-cause inpatient mortality rate of 4.16% (Table 3). Demographic trends were observed, particularly in the distribution of weight groups among race (Figures 1,2) and median household income (Figures 3,4). For both SHF and DHF, all-cause inpatient mortality was lowest in overweight patients followed by those with obesity and morbid obesity whereas mortality was highest in those with cachexia when compared to normal weight (Table 4).

**Figure 1:**
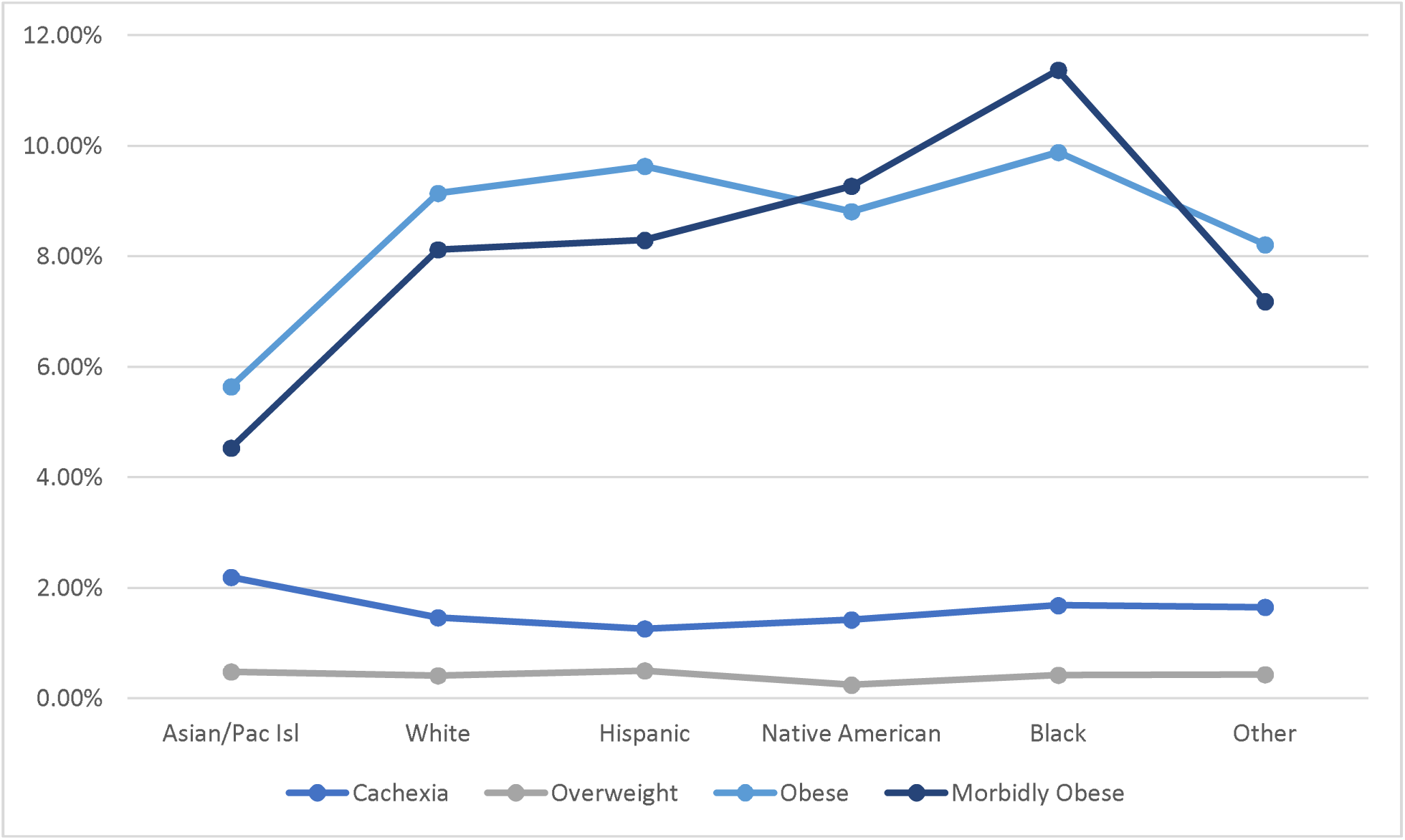
SHF demographic trends in the distribution of weight groups with race. Normal weight category omitted to better visualize trends of other groups.

**Figure 2:**
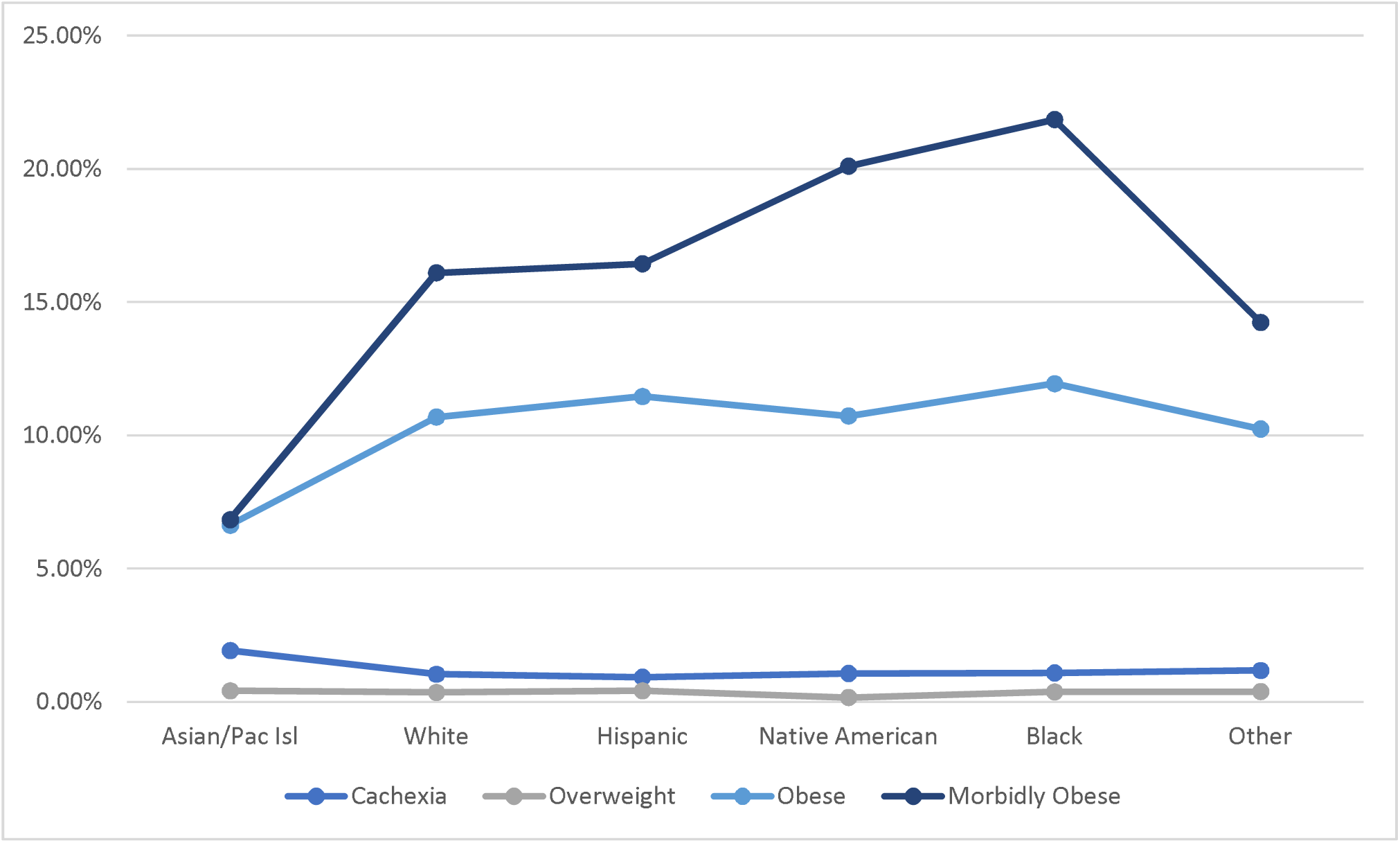
DHF demographic trends in the distribution of weight groups with race. Normal weight category omitted to better visualize trends of other groups.

**Figure 3:**
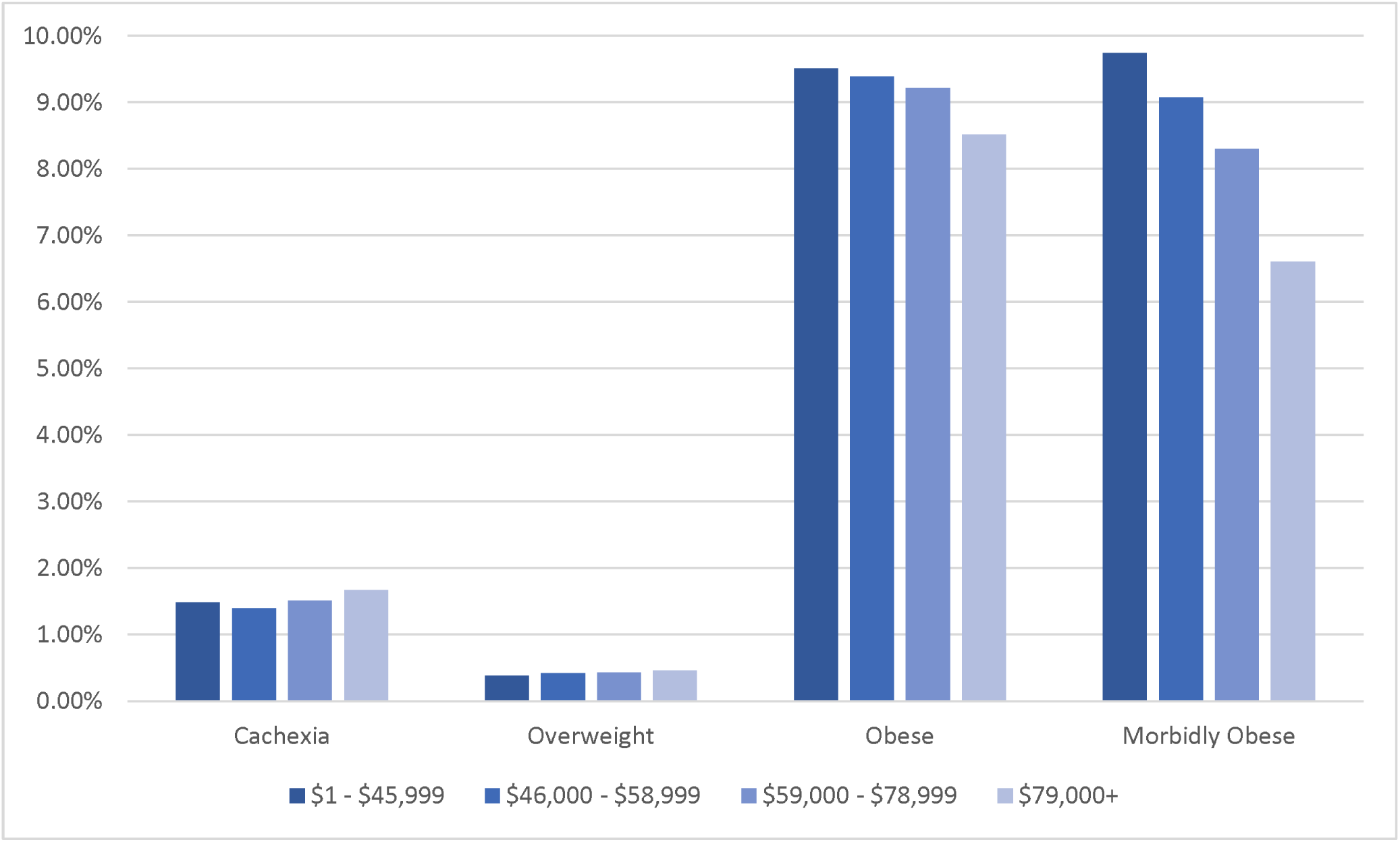
SHF demographic trends in the distribution of weight groups with median household income. Normal weight category omitted to better visualize trends of other groups.

**Figure 4:**
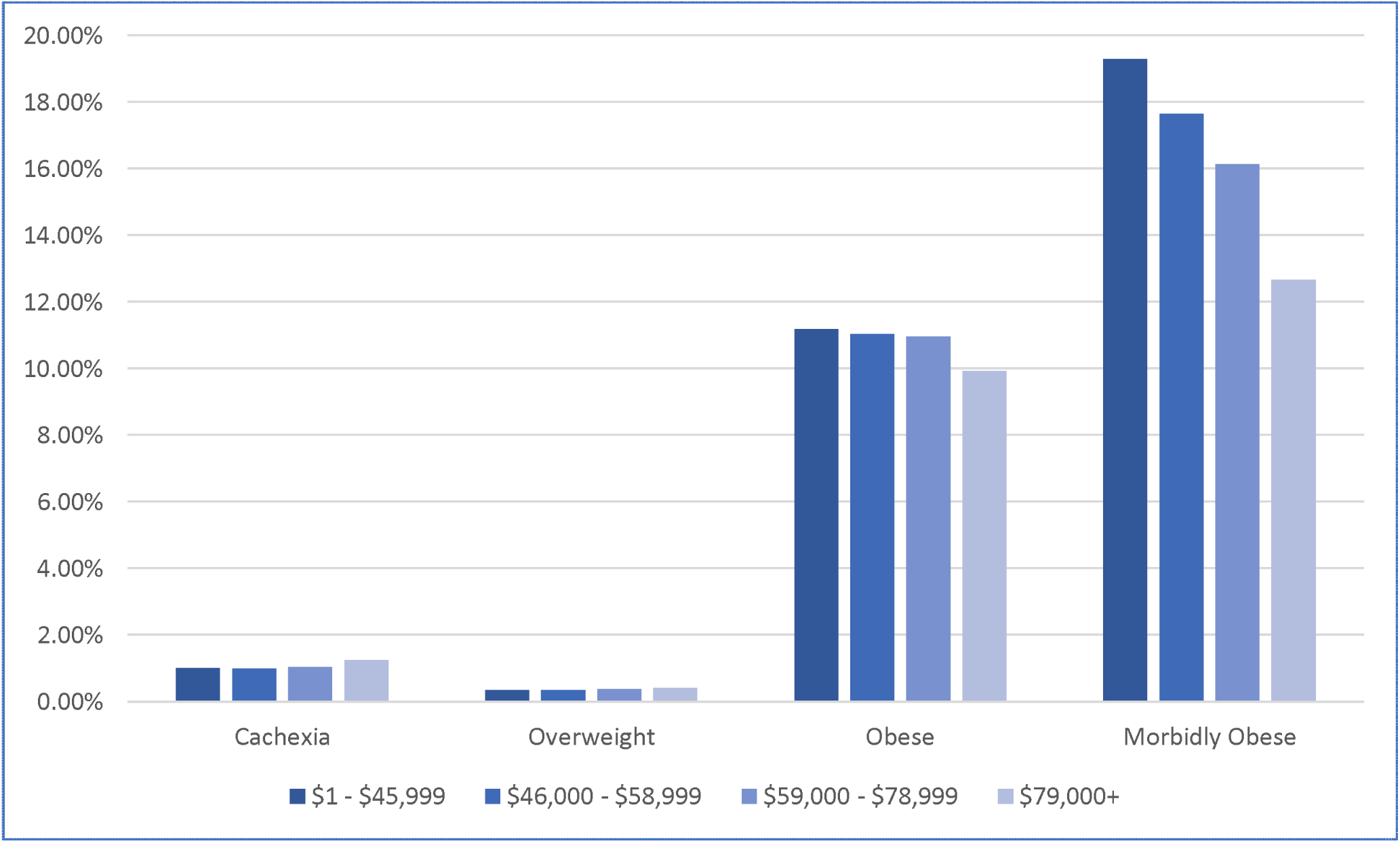
DHF demographic trends in the distribution of weight groups with median household income. Normal weight category omitted to better visualize trends of other groups.

**Table 2:** SHF patient demographics and details of their hospital visit (LOS = length of stay, SD = standard deviation, HMO = health maintenance organization).

|  |  |  |  |  |  |  |
| --- | --- | --- | --- | --- | --- | --- |
| 2016-2020, age >20 | Total SHF | Normal Weight | Cachexia | Overweight | Obesity | Morbid Obesity |
| Total Population | 7,364,023 | 5,911,238 | 110,755 | 30,640 | 679,350 | 639,765 |
| Age (Mean±SD) | 69.25±14.12 | 70.59±13.92 | 72.51±13.43 | 67.80±13.72 | 65.03±13.09 | 60.78±13.25 |
| LOS (Mean±SD) | 6±8 | 6±8 | 10±13 | 6±6 | 6±7 | 7±7 |
| Total Charges \$ (Mean±SD) | 84920±151322 | 84134±153412 | 114549±206703 | 81254±116750 | 85501±131882 | 86917±140842 |
| Mortality | 5.34% | 5.60% | 15.22% | 2.56% | 3.12% | 3.70% |
| Gender |  |  |  |  |  |  |
| Male | 63.76% | 81.01% | 1.48% | 0.44% | 9.21% | 7.95% |
| Female | 36.24% | 78.97% | 1.54% | 0.37% | 9.25% | 9.98% |
| Race |  |  |  |  |  |  |
| White | 66.49% | 80.97% | 1.46% | 0.41% | 9.14% | 8.12% |
| Black | 19.85% | 76.78% | 1.68% | 0.42% | 9.88% | 11.37% |
| Hispanic | 8.33% | 80.44% | 1.26% | 0.50% | 9.63% | 8.29% |
| Asian/Pacific Islander | 2.13% | 87.20% | 2.19% | 0.48% | 5.64% | 4.53% |
| Native American | 0.59% | 80.30% | 1.42% | 0.24% | 8.81% | 9.27% |
| Others | 2.61% | 82.64% | 1.65% | 0.43% | 8.21% | 7.18% |
| Primary Payer |  |  |  |  |  |  |
| Medicare | 69.09% | 82.64% | 1.66% | 0.39% | 8.23% | 7.18% |
| Medicaid | 11.97% | 75.11% | 1.36% | 0.48% | 10.41% | 12.78% |
| Private including HMO | 13.56% | 73.95% | 1.00% | 0.48% | 12.60% | 12.10% |
| Self-Pay | 2.89% | 75.65% | 0.90% | 0.43% | 11.41% | 11.74% |
| No Charge | 0.23% | 75.17% | 0.95% | 0.53% | 12.24% | 11.29% |
| Other | 2.26% | 79.49% | 1.39% | 0.62% | 10.02% | 8.57% |
| Hospital Bed size |  |  |  |  |  |  |
| Small | 18.57% | 80.18% | 1.39% | 0.46% | 9.16% | 8.92% |
| Medium | 28.19% | 80.46% | 1.42% | 0.40% | 9.01% | 8.82% |
| Large | 53.24% | 80.20% | 1.59% | 0.41% | 9.36% | 8.54% |
| Hospital Teaching/Location |  |  |  |  |  |  |
| Rural | 8.24% | 81.87% | 1.25% | 0.30% | 7.84% | 8.82% |
| Urban non-teaching | 19.82% | 80.60% | 1.21% | 0.42% | 8.74% | 9.13% |
| Urban Teaching | 71.94% | 80.00% | 1.61% | 0.43% | 8.52% | 8.55% |
| Hospital Region |  |  |  |  |  |  |
| Northeast | 19.01% | 82.26% | 1.89% | 0.48% | 8.34% | 7.10% |
| Midwest | 22.30% | 77.66% | 1.33% | 0.38% | 10.94% | 9.81% |
| South | 40.75% | 80.32% | 1.38% | 0.42% | 9.05% | 8.95% |
| West | 17.94% | 81.31% | 1.59% | 0.37% | 8.44% | 8.38% |
| Control/ownership of hospital |  |  |  |  |  |  |
| Government, nonfederal | 11.21% | 80.95% | 1.66% | 0.55% | 9.07% | 7.87% |
| Private, not-profit | 75.02% | 80.17% | 1.54% | 0.38% | 9.25% | 8.75% |
| Private, invest-own | 13.77% | 80.29% | 1.18% | 0.49% | 9.19% | 9.03% |
| Median Household Income |  |  |  |  |  |  |
| \$1 - \$45,999 | 33.89% | 79.01% | 1.49% | 0.38% | 9.51% | 9.74% |
| \$46,000 - \$58,999 | 26.73% | 79.84% | 1.40% | 0.42% | 9.39% | 9.07% |
| \$59,000 - \$78,999 | 22.42% | 80.63% | 1.51% | 0.43% | 9.22% | 8.30% |
| \$79,000+ | 16.96% | 82.82% | 1.67% | 0.46% | 8.52% | 6.61% |

**Table 3:** DHF patient demographics and details of their hospital visit.

|  |  |  |  |  |  |  |
| --- | --- | --- | --- | --- | --- | --- |
| 2016-2020, age >20 | Total DHF | Normal Weight | Cachexia | Overweight | Obesity | Morbid Obesity |
| Total Population | 10,064,223 | 7,152,273 | 106,815 | 37,230 | 1,091,110 | 1,694,205 |
| Age (Mean±SD) | 73.64±12.78 | 75.91±12.26 | 77.33±11.75 | 74.21±12.26 | 70.84±11.84 | 65.55±11.88 |
| LOS (Mean±SD) | 6±7 | 6±7 | 9±11 | 6±6 | 6±6 | 7±7 |
| Total Charges \$ (Mean±SD) | 70735±107650 | 69821±108281 | 98607±183102 | 68878±92710 | 69443±92654 | 73823±107626 |
| Mortality | 4.16% | 4.58% | 14.25% | 2.08% | 2.43% | 2.93% |
| Gender |  |  |  |  |  |  |
| Male | 41.02% | 72.08% | 1.03% | 0.41% | 11.13% | 15.51% |
| Female | 58.98% | 70.36% | 1.08% | 0.34% | 10.64% | 17.75% |
| Race |  |  |  |  |  |  |
| White | 72.63% | 71.97% | 1.04% | 0.36% | 10.69% | 16.10% |
| Black | 15.80% | 64.97% | 1.09% | 0.38% | 11.94% | 21.85% |
| Hispanic | 6.98% | 70.94% | 0.93% | 0.42% | 11.46% | 16.44% |
| Asian/Pacific Islander | 2.03% | 84.25% | 1.93% | 0.42% | 6.63% | 6.84% |
| Native American | 0.48% | 68.05% | 1.07% | 0.17% | 10.73% | 20.10% |
| Others | 2.08% | 74.11% | 1.18% | 0.39% | 10.24% | 14.24% |
| Primary Payer |  |  |  |  |  |  |
| Medicare | 79.84% | 73.66% | 1.12% | 0.37% | 10.39% | 14.61% |
| Medicaid | 7.32% | 57.59% | 0.89% | 0.35% | 11.82% | 29.59% |
| Private including HMO | 9.78% | 61.82% | 0.78% | 0.34% | 13.38% | 23.90% |
| Self-Pay | 1.40% | 61.72% | 0.74% | 0.37% | 12.64% | 24.77% |
| No Charge | 0.10% | 59.21% | 0.96% | 0.58% | 14.03% | 25.51% |
| Other | 1.56% | 68.85% | 0.95% | 0.42% | 11.55% | 18.41% |
| Hospital Bed size |  |  |  |  |  |  |
| Small | 21.12% | 71.00% | 0.99% | 0.39% | 10.85% | 16.93% |
| Medium | 29.51% | 71.18% | 1.03% | 0.37% | 10.67% | 16.92% |
| Large | 49.37% | 71.03% | 1.11% | 0.36% | 10.94% | 16.74% |
| Hospital Teaching/Location |  |  |  |  |  |  |
| Rural | 9.51% | 72.29% | 0.95% | 0.29% | 9.56% | 17.03% |
| Urban non-teaching | 20.91% | 71.52% | 0.97% | 0.38% | 10.17% | 17.14% |
| Urban Teaching | 69.58% | 70.76% | 1.10% | 0.38% | 11.22% | 16.72% |
| Hospital Region |  |  |  |  |  |  |
| Northeast | 20.47% | 74.39% | 1.26% | 0.49% | 10.08% | 13.90% |
| Midwest | 25.77% | 67.33% | 0.93% | 0.32% | 12.68% | 18.92% |
| South | 37.78% | 71.22% | 0.98% | 0.37% | 10.50% | 17.13% |
| West | 15.98% | 72.45% | 1.23% | 0.29% | 9.67% | 16.52% |
| Control/ownership of hospital |  |  |  |  |  |  |
| Government, nonfederal | 9.20% | 71.93% | 1.06% | 0.40% | 10.48% | 16.28% |
| Private, not-profit | 78.60% | 70.92% | 1.09% | 0.35% | 10.87% | 16.94% |
| Private, invest-own | 12.20% | 71.36% | 0.90% | 0.49% | 10.96% | 16.57% |
| Median Household Income |  |  |  |  |  |  |
| \$1 - \$45,999 | 30.27% | 68.36% | 1.01% | 0.35% | 11.19% | 19.29% |
| \$46,000 - \$58,999 | 26.85% | 70.14% | 1.00% | 0.35% | 11.04% | 17.64% |
| \$59,000 - \$78,999 | 23.87% | 71.65% | 1.04% | 0.38% | 10.96% | 16.14% |
| \$79,000+ | 19.01% | 75.88% | 1.25% | 0.41% | 9.92% | 12.67% |

**Table 4:** SHF and DHF all-cause inpatient mortality rates of each weight group compared to normal weight.

| **SHF** | Mortality | Normal weight<br>Mortality | Odds Ratio<br>(95% C.I.) | P-Value |
| --- | --- | --- | --- | --- |
| Cachexia | 15.22% | 5.60% | 3.02 (2.91-3.14) | <0.001 |
| Overweight | 2.56% | 5.60% | 0.44 (0.38-0.52) | <0.001 |
| Obesity | 3.12% | 5.60% | 0.54 (0.52-0.56) | <0.001 |
| Morbid<br>Obesity | 3.70% | 5.60% | 0.65 (0.63-0.67) | <0.001 |

| **DHF** | Mortality | Normal weight<br>Mortality | Odds Ratio<br>(95% C.I.) | P-Value |
| --- | --- | --- | --- | --- |
| Cachexia | 14.25% | 4.58% | 3.47 (3.32-3.61) | <0.001 |
| Overweight | 2.08% | 4.58% | 0.44 (0.38-0.52) | <0.001 |
| Obesity | 2.43% | 4.58% | 0.52 (0.50-0.53) | <0.001 |
| Morbid<br>Obesity | 2.93% | 4.58% | 0.63 (0.62-0.64) | <0.001 |

For both SHF and DHF, this relationship persists after multivariate analysis for comorbid conditions and baseline characteristics, as evident by the adjusted mortality odds ratios of each group in comparison to normal weight (Table 5). Mortality odds ratios of comorbidities in each group additionally show little variation in comparison to each other, indicating a lack of a confounding effect secondary to a comorbid condition (Figures 5,6).

**Figure 5:**
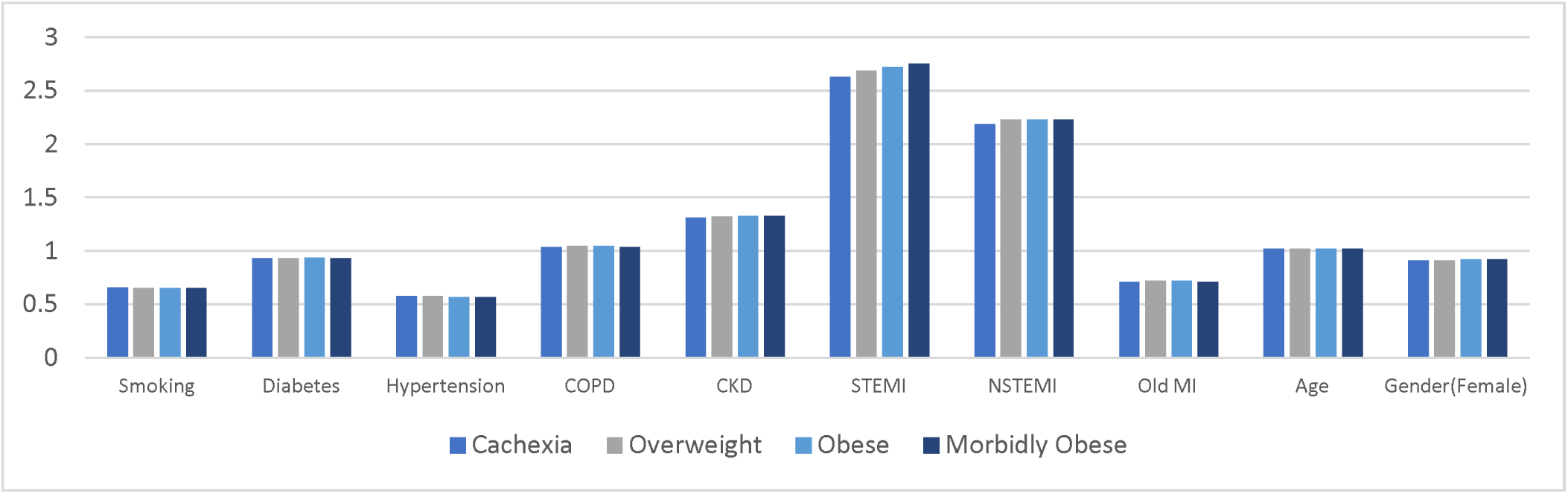
SHF mortality odds ratios of comorbidities.

**Figure 6:**
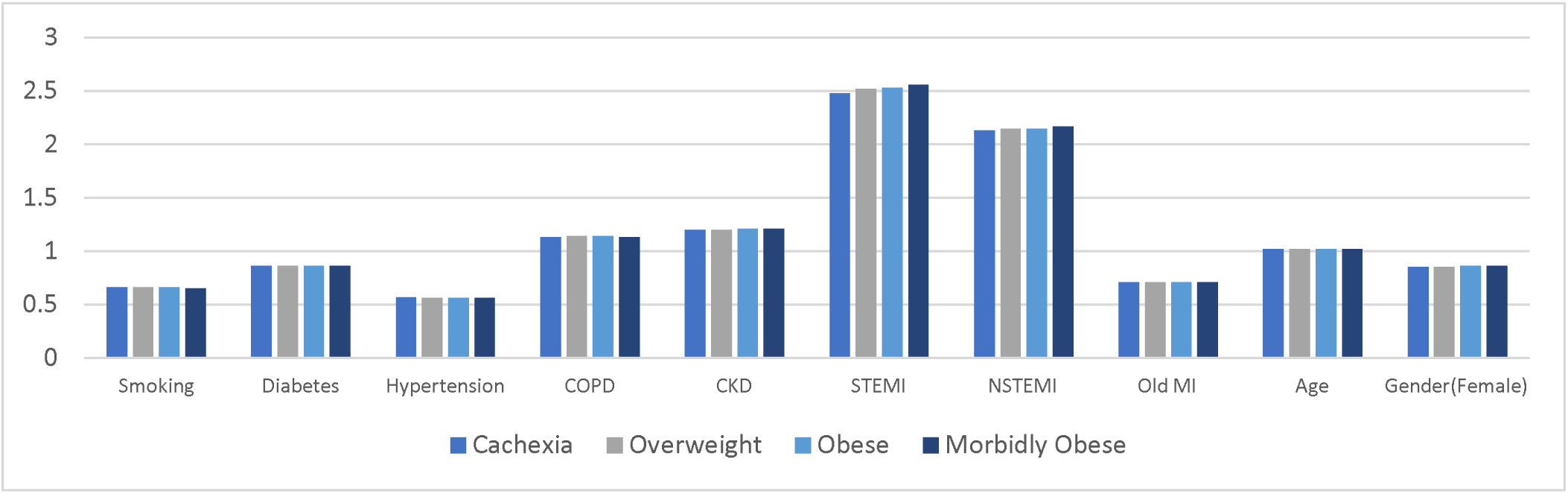
DHF mortality odds ratios of comorbidities.

**Table 5:** Multivariate SHF and DHF all-cause inpatient mortality odds ratios of each weight group in comparison to normal weight.

| **Multivariate SHF** | Odds Ratio<br>(95% C.I.) | P-Value | **Multivariate DHF** | Odds Ratio<br>(95% C.I.) | P-Value |
| --- | --- | --- | --- | --- | --- |
| Cachexia | 2.78 (2.67-2.90) | <0.001 | Cachexia | 3.09 (2.96-3.23) | <0.001 |
| Overweight | 0.49 (0.41-0.58) | <0.001 | Overweight | 0.47 (0.40-0.56) | <0.001 |
| Obesity | 0.64 (0.62-0.66) | <0.001 | Obesity | 0.61 (0.59-0.63) | <0.001 |
| Morbid Obesity | 0.85 (0.83-0.88) | <0.001 | Morbid Obesity | 0.83 (0.81-0.85) | <0.001 |

## Discussion

Cachexia is a devastating illness that clinically presents as progressive weight loss with alterations in body composition and disturbed homeostasis of several body systems. Particularly, cachexia is characterized by the loss of muscle mass, which may or may not be accompanied by a loss of fat mass. It can manifest in a variety of diseases, including HF (termed cardiac cachexia in this context), and is related to increased morbidity and mortality.^14,15^ Our findings support this and it reinforces the external validity of our study, but more importantly, cardiac cachexia may provide an explanation for the underlying physiology of the obesity paradox in CVD. It has been shown that in the context of altered levels of endocrine mediators such as insulin, insulin-like growth factor 1, leptin, ghrelin, melanocortin, growth hormone, and neuropeptide Y that lead to cardiac cachexia, cardiac obesity actually plays a protective role.^14,15^ That is, cardiac obesity is thought to mitigate the increased morbidity and mortality that would normally be witnessed in these patients secondary to cardiac cachexia. Furthermore, adipose tissue has been shown to be beneficial to cardiac cachexia in more than one way. It has been recently demonstrated that in those with cachexia and lower fat/lean body mass ratio, there is more right ventricle (RV) dysfunction, suggesting that increased adipose tissue relative to lean mass may be cardioprotective in the context of HF.^6,16^ It follows that those who have a higher fat/lean body mass ratio may develop cardiac cachexia less, and have lower mortality rates in HF. This further highlights the paradoxical and multifaceted cardioprotection that excess fat provides in the context of HF, as it mitigates cardiac cachexia which is linked to significantly increased mortality in these patients.

Our data is consistent with these hypotheses and displays the weight at which excess adiposity is the most cardioprotective while still minimizing the detrimental effects of obesity. That is, HF patients with less adiposity than the overweight HF patients do not reap the benefit of cardioprotection, but those with more adiposity than the overweight group face the detrimental effects that excess adipose tissue exerts on their bodies. Still, HF patients who are obese and morbidly obese have lower all-cause inpatient mortality rates than those of normal weight, suggesting that the benefit of cardioprotection outweighs the cost of the negative metabolic effects of excess adiposity on the body in these populations.

Conversely, some have hypothesized that the patterns observed in the obesity paradox may not be secondary to excess fat mass playing a protective role, but instead are related to levels of lean mass.^5,6,13,17,18^ In this context, lean refers to fat-free mass, such as skeletal muscle. A deficiency in lean mass, also known as sarcopenia, is independently associated with a poor prognosis in numerous chronic diseases, including HF.^19,20^ If this was the case, findings would likely manifest with patients of lower BMI having worse outcomes in comparison to those with higher BMI. This may be due to the relationship between skeletal muscle mass and cardiorespiratory fitness (CRF),^13,21^ defined by the minute ventilation divided by carbon dioxide production (VLJ_E_ / VLJ_CO2_) slope. This is a measure that quantifies ventilatory efficiency and a lower VLJ_E_ / VLJ_CO2_slope has been shown to correlate with better outcomes in HF patients, irrespective of body habitus.^13^ It follows that higher amounts of lean mass exert protective effects on patients with HF, and/or the detrimental effects of sarcopenia manifest as increased mortality in lower BMI categories. This also suggests that among two HF patients of similar weights in the obese category of BMI, with one being nonsarcopenic obese and the other being sarcopenic obese, the former would have higher CRF and better outcomes.^13^

Our data is also consistent with this idea of skeletal muscle’s role in CRF, and further ties in with the idea that cardiac cachexia plays a significant role in HF mortality. Still, these competing hypotheses function on different ideologies regarding the underlying physiology of the obesity paradox. Is it increased fat mass, increased lean mass, or a combination of the two that are protective from mortality in HF? This question sheds light on the primary limitation of those studying obesity in all fields of research, including our own HF study. We used ICD-10 diagnosis codes, which function on a basis of BMI, to identify and categorize our patients. BMI is a measure that has widespread standardized use in healthcare, and therefore utilizing it has allowed us to identify and categorize an extremely large number of HF patients. However, this comes at a cost since BMI does not distinguish between variations in body composition (fat mass versus lean mass) and fat distribution (subcutaneous fat versus visceral fat).^6,17,18,22^ As a result, there may be patients with different body compositions and more importantly, different metabolic profiles, who get placed into the same BMI group. BMI is an incredibly quick, easy, and useful tool for healthcare workers to get a snapshot of a patient’s potential body type and allows researchers to have an abundance of data to analyze. However, due to the variation in body composition within BMI groups, it has been argued that there are more effective measures of adiposity such as waist circumference, waist-to-hip ratio, body fat percentage, and fat mass index.^3,6,23^ These alternative measures are not nearly as commonly used in medicine as BMI is. Furthermore, they require more time with patients, training of healthcare workers, and increased cost of visits.^5^ As such, this may introduce challenges to those studying obesity in any field, as most patients obtain a BMI measurement when they are receiving healthcare, and very few obtain a measurement with one of the alternative techniques mentioned above. Even more so, it may be warranted to obtain measures of lean mass alongside these alternative measures of fat mass to obtain a more complete picture of the underlying physiology of the obesity paradox.

Although some have criticized the use of BMI as a predictor of mortality in CVD, it has been demonstrated that at the population level, BMI still predicts clinical outcomes and that BMI can be as clinically important as the other total adiposity measures.^24^ In fact, it has also been shown that these other measures of fat mass still demonstrate the obesity paradox.^5^ Given these findings, BMI will likely continue to be the gold standard of body composition assessment due to its utility, simplicity, widespread adoption in literature and healthcare, and endorsement by groups such as the World Health Organization.^2,24^ Overall, in our own study, we have demonstrated a strong pattern of inpatient mortality rates in HF patients of varying BMI, but we are still unable to say with confidence what the underlying physiology of the obesity paradox is to explain our findings. Now that the pattern of the obesity paradox has been mapped out by the largest study to date, further prospective studies with other measures of adiposity and lean mass are warranted to analyze the physiological relationship of HF and obesity and distinguish which metabolic processes are at play to generate these patterns. Obesity is an incredibly complex disease, and the pattern witnessed in the obesity paradox is likely multifaceted with physiology that cannot be explained by a single mechanism.

It should be noted that although we have confirmed and further characterized the obesity paradox in SHF and DHF, this should not be interpreted as a reason to promote habits leading to overweight or obese individuals in any population. Since obesity is a significant risk factor in the pathogenesis of many chronic health conditions, including CVD and specifically HF, individuals who endorse healthy habits are practicing primary prevention of the development of HF in the first place.

Lastly, by analyzing the demographics of our study, particularly race and median household income, we observe a clear example of the social determinants of health and take this opportunity to advocate for more equitable healthcare. It is apparent that White and Asian/Pacific Islander populations make up a larger proportion of the normal weight category, whereas those who are Black or Native American and make up a much larger proportion of the obese and morbidly obese categories. This is the pattern we expect to see,^25,26,27,28^ serving as a source of external validity of our study, but more importantly highlights the importance of equitable healthcare. It is critical to be mindful of how these communities are disproportionately affected by systemic issues, such as limited access to healthcare, less educational opportunities, food deserts, and decreased economic stability,^1,25,27^ which all lead to a worsened overall health of an individual. An expected pattern is also witnessed with median household income. We see a strong inverse relationship between median household income and rates of obesity and morbid obesity. This further emphasizes just how much of a role that economic stability and healthcare access play in an individual’s overall health. It is essential that healthcare workers and researchers alike continue to advocate for more equitable healthcare to address these disparities.

## Conclusion

Our data concludes that after multivariate adjustment for comorbidities, all-cause inpatient mortality in SHF and DHF is lowest in overweight patients. Obese and morbidly obese SHF and DHF patients also have lower all-cause inpatient mortality compared to normal-weight patients but to a lesser extent than those that are overweight. Although we confirmed the obesity paradox in HF patients, we found that increasing weight above the overweight threshold reduces the beneficial effect of additional weight in this population. Patients with cachexia by far have the highest all-cause inpatient mortality, nearly tripling the mortality rates of HF patients of normal weight. Further prospective studies with other measures of adiposity and lean mass are warranted to analyze the physiological relationship of HF and obesity and distinguish which metabolic processes are at play to generate these patterns.

### Limitations

We used administrative ICD-10 coding with inherent limitations. Our study was a retrospective study needing confirmation in a prospective study. We adjusted for many comorbidities but we can not rule out other important comorbidities that could lead to obesity paradox not adjusted in our study.

## Data Availability

NIS data base publically available

## References

1. Roberto CA, Swinburn B, Hawkes C, et al. Patchy progress on obesity prevention: emerging examples, entrenched barriers, and new thinking. The Lancet. 2015;385(9985):2400-2409. doi:10.1016/S0140-6736(14)61744-X

2. Boutari C, Mantzoros CS. A 2022 update on the epidemiology of obesity and a call to action: as its twin COVID-19 pandemic appears to be receding, the obesity and dysmetabolism pandemic continues to rage on. Metabolism. 2022;133:155217. doi:10.1016/j.metabol.2022.155217

3. Mathieu P, Poirier P, Pibarot P, Lemieux I, Després JP. Visceral obesity: the link among inflammation, hypertension, and cardiovascular disease. Hypertension. 2009;53(4):577–584. doi:10.1161/HYPERTENSIONAHA.108.110320

4. Kawai T, Autieri MV, Scalia R. Adipose tissue inflammation and metabolic dysfunction in obesity. American Journal of Physiology-Cell Physiology. 2021;320(3):C375–C391. doi:10.1152/ajpcell.00379.2020

5. Elagizi A, Kachur S, Lavie CJ, et al. An overview and update on obesity and the obesity paradox in cardiovascular diseases. Progress in Cardiovascular Diseases. 2018;61(2):142–150. doi:10.1016/j.pcad.2018.07.003

6. Clark AL, Fonarow GC, Horwich TB. Obesity and the obesity paradox in heart failure. Progress in Cardiovascular Diseases. 2014;56(4):409–414. doi:10.1016/j.pcad.2013.10.004

7. Lavie CJ, De Schutter A, Parto P, et al. Obesity and prevalence of cardiovascular diseases and prognosis—the obesity paradox updated. Progress in Cardiovascular Diseases. 2016;58(5):537–547. doi:10.1016/j.pcad.2016.01.008

8. Davos CH, Doehner W, Rauchhaus M, et al. Body mass and survival in patients with chronic heart failure without cachexia: The importance of obesity. Journal of Cardiac Failure. 2003;9(1):29–35. doi:10.1054/jcaf.2003.4

9. Horwich TB, Fonarow GC, Hamilton MA, MacLellan WR, Woo MA, Tillisch JH. The relationship between obesity and mortality in patients with heart failure. Journal of the American College of Cardiology. 2001;38(3):789–795. doi:10.1016/S0735-1097(01)01448-6

10. Shah R, Gayat E, Januzzi JL, et al. Body mass index and mortality in acutely decompensated heart failure across the world. Journal of the American College of Cardiology. 2014;63(8):778–785. doi:10.1016/j.jacc.2013.09.072

11. Kenchaiah S, Pocock SJ, Wang D, et al. Body mass index and prognosis in patients with chronic heart failure: insights from the candesartan in heart failure: assessment of reduction in mortality and morbidity (Charm) program. Circulation. 2007;116(6):627–636. doi:10.1161/CIRCULATIONAHA.106.679779

12. Fonarow GC, Srikanthan P, Costanzo MR, Cintron GB, Lopatin M. An obesity paradox in acute heart failure: Analysis of body mass index and inhospital mortality for 108927 patients in the Acute Decompensated Heart Failure National Registry. American Heart Journal. 2007;153(1):74–81. doi:10.1016/j.ahj.2006.09.007

13. Ortega FB, Lavie CJ, Blair SN. Obesity and cardiovascular disease. Circ Res. 2016;118(11):1752-1770. doi:10.1161/CIRCRESAHA.115.306883

14. Soto ME, Pérez-Torres I, Rubio-Ruiz ME, Manzano-Pech L, Guarner-Lans V. Interconnection between cardiac cachexia and heart failure—protective role of cardiac obesity. Cells. 2022;11(6):1039. doi:10.3390/cells11061039

15. Selthofer-Relatić K, Kibel A, Delić-Brkljačić D, Bošnjak I. Cardiac obesity and cardiac cachexia: is there a pathophysiological link? Journal of Obesity. 2019;2019:1–7. doi:10.1155/2019/9854085

16. Melenovsky V, Kotrc M, Borlaug BA, et al. Relationships between right ventricular function, body composition, and prognosis in advanced heart failure. Journal of the American College of Cardiology. 2013;62(18):1660–1670. doi:10.1016/j.jacc.2013.06.046

17. Romero-Corral A, Somers VK, Sierra-Johnson J, et al. Diagnostic performance of body mass index to detect obesity in patients with coronary artery disease. European Heart Journal. 2007;28(17):2087–2093. doi:10.1093/eurheartj/ehm243

18. Carbone S, Lavie CJ, Arena R. Obesity and heart failure: focus on the obesity paradox. Mayo Clinic Proceedings. 2017;92(2):266–279. doi:10.1016/j.mayocp.2016.11.001

19. Von Haehling S, Morley JE, Anker SD. An overview of sarcopenia: facts and numbers on prevalence and clinical impact. J cachexia sarcopenia muscle. 2010;1(2):129–133. doi:10.1007/s13539-010-0014-2

20. Bekfani T, Pellicori P, Morris DA, et al. Sarcopenia in patients with heart failure with preserved ejection fraction: Impact on muscle strength, exercise capacity and quality of life. International Journal of Cardiology. 2016;222:41–46. doi:10.1016/j.ijcard.2016.07.135

21. Balady GJ, Arena R, Sietsema K, et al. Clinician’s guide to cardiopulmonary exercise testing in adults: a scientific statement from the american heart association. Circulation. 2010;122(2):191–225. doi:10.1161/CIR.0b013e3181e52e69

22. Romero-Corral A, Somers VK, Sierra-Johnson J, et al. Accuracy of body mass index in diagnosing obesity in the adult general population. Int J Obes. 2008;32(6):959–966. doi:10.1038/ijo.2008.11

23. Kuriyan R. Body composition techniques. Indian J Med Res. 2018;148(5):648. doi:10.4103/ijmr.IJMR_1777_18

24. Ortega FB, Sui X, Lavie CJ, Blair SN. Body mass index, the most widely used but also widely criticized index. Mayo Clinic Proceedings. 2016;91(4):443–455. doi:10.1016/j.mayocp.2016.01.008

25. Heymsfield SB, Peterson CM, Thomas DM, Heo M, Schuna JM. Why are there race/ethnic differences in adult body mass index–adiposity relationships? A quantitative critical review. Obesity Reviews. 2016;17(3):262–275. doi:10.1111/obr.12358

26. Wagner DR, Heyward VH. Measures of body composition in blacks and whites: a comparative review. The American Journal of Clinical Nutrition. 2000;71(6):1392–1402. doi:10.1093/ajcn/71.6.1392

27. Deurenberg P, Yap M, Van Staveren W. Body mass index and percent body fat: a meta analysis among different ethnic groups. Int J Obes. 1998;22(12):1164–1171. doi:10.1038/sj.ijo.0800741

28. Hsu WC, Boyko EJ, Fujimoto WY, et al. Pathophysiologic differences among asians, native hawaiians, and other pacific islanders and treatment implications. Diabetes Care. 2012;35(5):1189–1198. doi:10.2337/dc12-0212

